# A Comprehensive Analysis of COVID-19 Outbreak situation in India

**DOI:** 10.1101/2020.04.08.20058347

**Authors:** Rajan Gupta, Saibal K. Pal, Gaurav Pandey

## Abstract

The outbreak of COVID-19 in different parts of the world is a major concern for all the administrative units of respective countries. India is also facing this very tough task for controlling the virus outbreak and has managed its growth rate through some strict measures. This study presents the current situation of coronavirus spread in India along with the impact of various measures taken for it. With the help of data sources (till 7^th^-8^th^ April 2020) from various state units of India and Ministry of Health and Family Welfare, Government of India, this study presents various trends and patterns. This study answers six different research questions in a comprehensive manner. It has been reported that growth rate of infected cases has been controlled with the help of National Lockdown, however some uncontrolled mass level events had a negative impact on the infected cases. With the help of exponential and polynomial regression modelling, the predictions of up to 75000 cases have been done by the end of April 2020. It has also been seen that there are some prominent clusters and patient nodes in the network of patients which are the major influencers for COVID-19 spread. Also, death rate case predictions have been done through two-class classification models with an accuracy of 60%. At the end, strategies for continuation for lockdown has been discussed and presented. It appears that only essential services should be open for the citizens of India and the national lockdown should be carried on for next 2-4 weeks. This study will be useful for the Government of India and various states of India, Administrative Units of India, Frontline health workforce of India, researchers and scientists. This study will also be favorable for the administrative units of other countries to consider various aspects related to the control of COVID-19 outspread in their respective regions.

## 1 Introduction

COVID-19, or more popularly known as Novel Corona Virus, is associated with the respiratory disorder in humans which has been declared as a global epidemic and pandemic in the first quarter of the year 2020 by the World Health Organization [1]. As per the latest data (7^th^ April 2020) by John Hopkins University [2] and other tracking websites, there are currently more than 1.3 million people infected by the Novel Corona Virus all around the world and close to 75 thousand deaths reported from different parts of the world. The top 10 countries with maximum number of infected cases are the United States of America, Spain, Italy, Germany, France, China, Iran, United Kingdom, Turkey and Switzerland. The top countries with maximum number of reported deaths are Italy, Spain, United States of America, France and United Kingdom. With respect to the recovered patients list, China is at the top of the list followed by Spain, Germany, Italy, Iran and the United States of America. India (https://www.mohfw.gov.in/) was placed comfortably out the list of infected nations by huge margins, but recent events led to its rise to 27^th^ position (on 7^th^ April 2020) which is a point of concern. The mortality rate is controlled at less than 3% right now, which is better than the ∼5.5% mortality rate of world, but the model of spread is slowly moving towards an exponential trend which can lead to massive loss of lives and infrastructure (https://www.mygov.in/covid-19/).

India is being looked upon by various nations now as a World Leader and even WHO acknowledged that world is looking towards Indian strategies to contain the outbreak of this epidemic [3]. India accounts for almost one-fifth of the world’s population and is second leading country in terms of population in the world. India contributes heavily to the world’s GDP and is amongst the most prominent developing country in the world with fairly strong economic growth percentages [4]. India’s good camaraderie with majority of the nations in the world and its helpful nature makes it a perfect ally for other countries. Therefore, the analysis of COVID-19 outbreak in Indian region is closely watched and monitored by the World and there is a need of comprehensive analytical studies based on different strategies taken by Indian administrators from time to time. India has been following a nationwide lockdown since 22-March-2020, which was a one-day lockdown, followed by a 21-day lockdown after two days. Every activity in India since then has been happening with the permission from various administration units and almost all the domestic and international travels have been either banned or monitored closely. India is yet to get into the third phase of COVID-19 outbreak i.e. the community outbreak as seen by various countries around the world, but the cases have been rising continuously. India’s lockdown period has been impacted by two major events in the recent days which were related to the mass exodus of laborers and workers from one state to other states (especially from Delhi to neighboring states) and conduction of a religious event in Delhi which led to spike in the number of cases in various states of India. During this time, the Indian Prime Minister has been trying to connect with Indian citizens through innovative strategies and coming up with various engagement activities which are impacting the whole nation. With so much happening in India right now, it becomes imperative that we study the current situation and impact of various such events in India through data analysis methods and come up with different plans for future which can be helpful for the Indian administrators and medical professionals.

The current study explores various aspects associated with the COVID-19 outbreak in India and the various regions situated in India. The specific research questions (RQ) explored in this study are as follows.

- RQ1: How has the situation changed in post-lockdown period in India i.e. what is the outbreak situation after 22-March-2020 in India as compared to pre-lockdown period?
- RQ2: What are the short terms predictions for the number of infected cases in India for the next 3-4 weeks based on current situation?
- RQ3: Has the lockdown been followed by the Indian citizens after 22-March-2020? Has the Social Distancing worked for Indian citizens? What are the mobility changes in the various regions of India?
- RQ4: Whether the community outbreak spread started in India with the conduct of a religious event in Delhi? How is the outbreak different for citizens related to event and for citizens not related to the event?
- RQ5: Which are the prominent clusters which were formed in the last few weeks with respect to COVID-19 outbreak in India? Does Network Analysis provide influential points in the infected patient network?
- RQ6: Whether the national lockdown should be opened after 14^th^ April 2020 in India or should it continue? Are there any partial regions for which lockdown can be removed? Which all essential services should be opened in India after one week under restricted lockdown?

The current study is divided into five sections. First section has laid the context of the study. The second section discusses various literature review and analytical techniques followed by the different researchers across the world. The third section presents the methodology and research variables of the study. The fourth section presents the results and findings of the study along with the discussion of the achievement of the various research questions explored in this study. And finally section five concludes this study and present limitations and future directions for this research work.

## 2 Literature Review

As per different papers available in literature, there are a few studies that focus on the trend analysis and forecasting for Indian region. The studies [5][6] on Indian region presents long term and short term trend, respectively. These studies used time series data from John Hopkins University database and presented forecasting using ARIMA model, Exponential Smoothing methods, SEIR model and Regression Model. However network modeling and pattern mining were not attempted in these versions of the studies and that too at the regional level, hence the current study attempts to do that. Also, the studies in Indian region from the past are more focused on presenting time series analysis based on the overall data for Indian region rather than covering other sources of information apart from just considering the number of infected patients, so the need to analyze the patients background and information is required for the authorities to get better insight about the situation.

Similarly, there are other mathematical models that were developed for analyzing the trends of COVID-19 outbreak in India. A model [7] for studying the impact of social distancing on the age and gender of the patients in India was presented. It compared the country demographics amongst India, Italy and China and suggested the most vulnerable age categories and gender groups amongst all the nations. The study also predicted the rise of infected cases in India with different lockdown periods. Similarly, a network structure approach was used by one of the study [8] to see whether any specific node clusters were getting formed. But only travel data nodes were considered by the authors to check which the prominent regions are affecting Indian travelers coming back to the India. Also, the study presented the SIR model to see the rate of spread of the Corona Virus amongst patients in India. Analysis on the testing labs and infrastructure was also presented by earlier authors.

Work of medical practitioners and frontline health workers was also presented by some studies [9]. It was found that in India, the role of health workers was less stressed as the spread stage of corona virus was still in phase two or the phase of local transmission rather than the community transmission as compared to other nations like Italy, Spain and USA. However, it was also claimed that Indian healthcare infrastructure is not very strong as per the WHO guidelines and in case of community spread, the Indian government may find it difficult to manage the spread. Some detailed discussion on the nature of the Corona Virus was also presented by some studies [10][11].

Apart from India, a few models are also available for other countries, primarily for China, Italy and USA, as the numbers of infected patients are high. Studies like [12-16] worked on various mathematical models to determine the spread of the disease, predict the number of infected patients, commenting on the preparedness for each country in tackling COVID-19 spread and finding the patterns of flattening curve in different conditions. A lot of researches are still in preprint stage for the world level and are yet to be peer reviewed.

With respect to the research activities conducted in the Indian region, the studies are yet to work on the impact of different policies working towards containment of the corona virus. Even in the preprint databases, there are fewer evidences available which worked in the Indian region with more granularities and came up with analysis that can support the decision making of the various administrators in India to curb the lockdown and work on future strategies. Therefore, this study attempts to work on a comprehensive level to analyze the COVID-19 spread in India and impact of various strategies imposed by the Government at both state level and central level.

## 3 Methodology

To answer the different research questions, specific methodologies have been used which covers up different data set, data sources, modeling techniques and outcome variables. The overall variables covered up in the study are shown in Figure 1.

**Figure 1.**
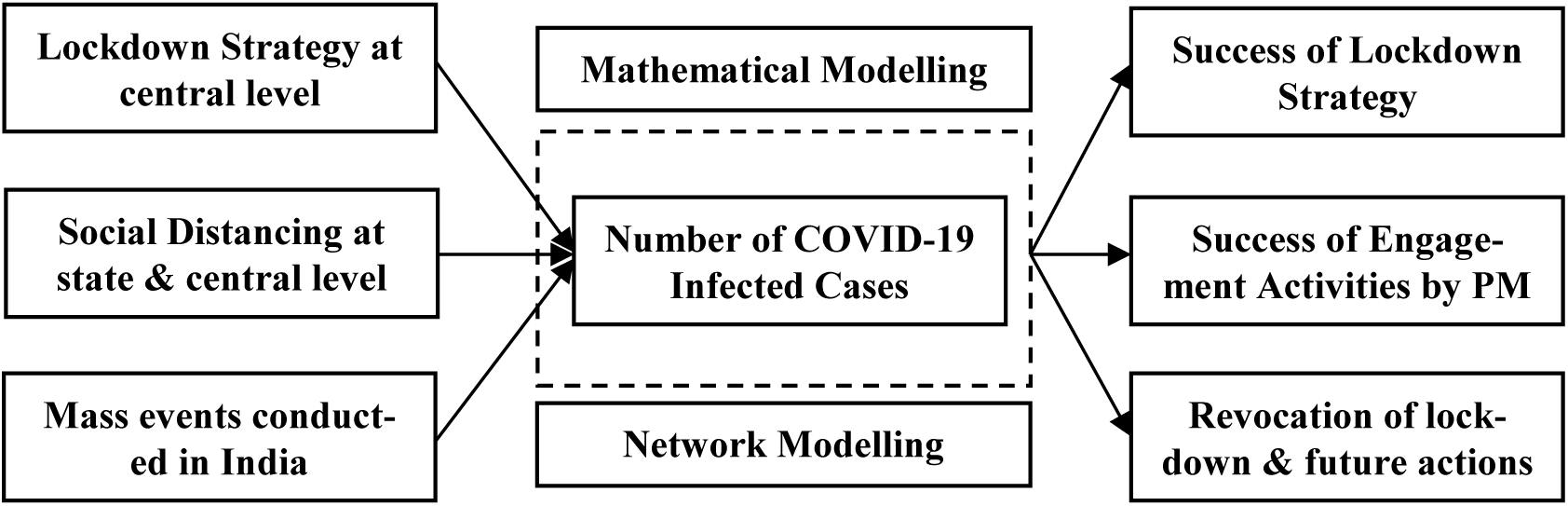
Research Model for the current study covering various variables and outcome measures.

For the first research question, the time series data from the time period of 30^th^ January 2020 till 7^th^ April 2020 has been covered from the Indian database of COVID-19 [17]. This dataset was divided into three parts of 30^th^ January 2020 to 4^th^ March 2020, 5^th^ March to 22^nd^ March 2020 and 23^rd^ March 2020 to 7^th^ April 2020. Trend analysis and average number of infected cases were compared at both the national level. On the same dataset, the second research question has been answered using the Exponential Modelling to predict the short term trends for the next three weeks. The exponential modelling can be done using equation 1.

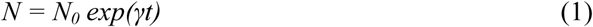

Where, N_0_ and N are the total cases at time t_0_ and t, respectively, and γ is the growth rate. By definition [18], N = 2N_0_ needs the doubling time (α) that is expressed as *α = ln(2)/γ* through *ln(N/N*_*0*_*) = ln(2) = γt*. If the growth rate evolves with time, depending on the effective prevention strategies, then the doubling time changes with time. Thus, the time-dependent growth rate *γ(t)* causes the time-dependent doubling time *α(t)*, defined as *α(t) = t ln(2)/ ln(N/N*_*0*_) through *γ(t) = ln(N/N*_*0*_*)/t*.

For the third research question, the mobility data set [19] from Google has been considered after 22^nd^ March 2020 to see the patterns in mobility of Indian citizens at prominent places in India. Also, the number of operational places and events conducted during lockdown data has been analyzed through news reports and channels. Based on the news reports, the major events were traced and people getting infected due to such events have been covered from the Indian data repository created using crowdsourcing channels [20]. The patients were categorized as infected from the event as compared to other patients who were not related to the event. The trends and patterns were analyzed after conduction of this event i.e. in the first week of April 2020. The data was also confirmed from various news reports in India.

To understand the patterns existing in the patients of COVID-19 in India, their demographic details were studied in detail since 30^th^ January 2020 from the crowdsourced database in India [20] and rules were generated for the patients with potential probabilities. Network Analysis [21] was also conducted on the patients and node centralities [22] were calculated for the patients. Based on the patient database and number of infected cases along with the various secondary reports from the news media and consulting firms, the success of engagement activities and lockdown strategies have been analyzed, which answers the 6^th^ research question.

## 4 Findings and Discussion

The findings of the different types of data analysis conducted on various types of datasets are presented in the current section as follows.

### a. Impact of Lockdown on Infected cases

The national lockdown for one day was announced on 22^nd^ March 2020 by the central government of India, before which majority of the schools, colleges, markets, cinema halls, etc. were already shut down by respective state governments. Merely, two days after this one day curfew, a 21-day lockdown was announced by the central government banning all the movement and restricting the Indians to stay at home. The citizens were allowed to step out of their homes only in the emergency situations and that too with prior permissions from the local administration. All these instructions were given in the hope of flattening the curve of infected cases and to restrict the exponential growth of the patients in India.

There are almost 5000 confirmed cases reported in India as on the morning of 7^th^ April 2020 with more than 90% cases being active. The death rate has been keeping under 3% at all the stages of the COVID-19 spread. Looking at the graph in Figure 2, it is clearly evident that a spike has been reported in India after 22^nd^ March 2020 i.e. the time when lockdown was announced. It clearly shows that Indian authorities were quick enough to sense the spread rate in Indian region and taking necessary steps of maintaining social distancing by announcing a rigid step of lockdown.

**Figure 2.**
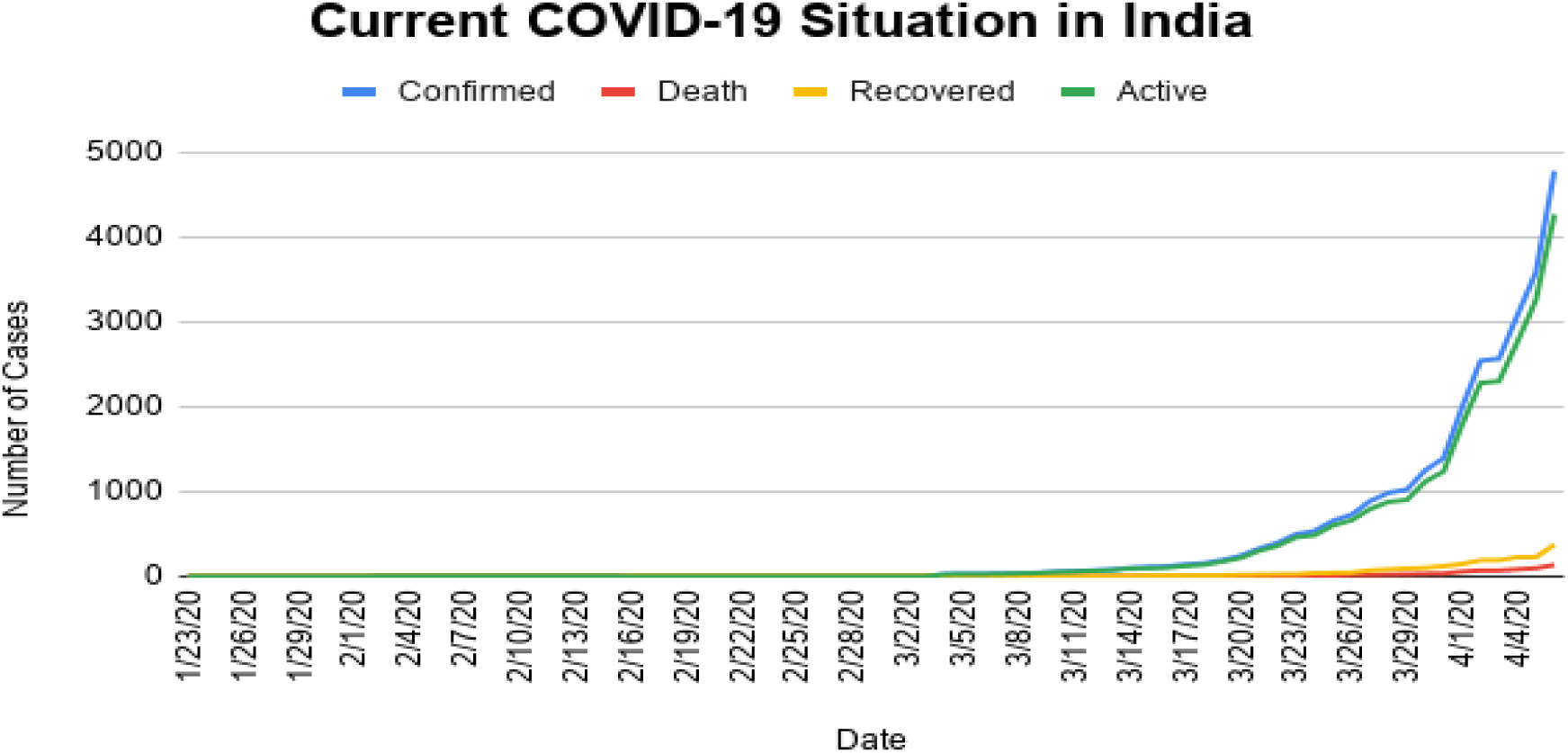
The number of COVID-19 cases in India till 6^th^ April 2020.

Looking at the growth rate of infected cases on daily basis (blue line) and the trend line of expected growth (red line) in Figure 3, it appears that in early days of infection the growth rate was quite high due to low number of cases. The growth rate has been calculated as the difference in number of cases between two consecutive days, divided by the count of infected cases on the previous day of the two days under consideration, multiplied by 100. Since, in the early days the count was in single digits, so the growth rate was pretty high. Hence ignoring that period, considering the second phase of time period from 5^th^ March to 22^nd^ March 2020 i.e. exactly before the lockdown, the growth rate has been hovering around 20% with trend line forecasting it to be maximum around 28%. However, for the time period after lock-down i.e. from 22^nd^ March 2020 onwards, the growth rate slightly increased, but it remained around a similar mark of 20%. And the trend line also predicted the growth rate on per day basis to be around maximum of 28%. Therefore, it can be said that the national lockdown has been able to contain the growth of the number of cases of COVID-19 patients. Without lock-down, the growth might not have been contained in India and may have gone into the exponential zone too quickly. This gives all the state level and national level administrators and health workers to get prepared for the rising number of cases.

**Figure 3.**
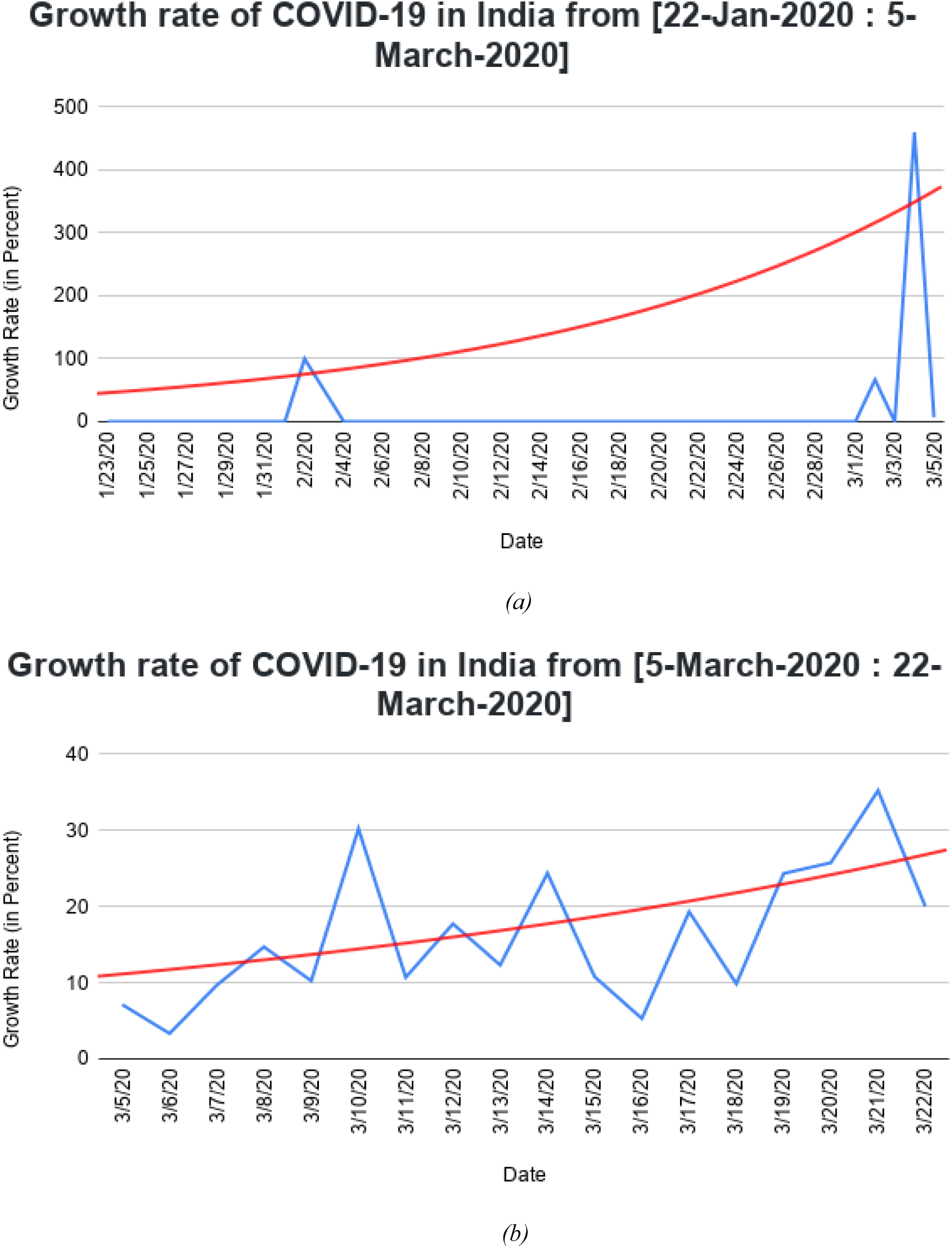
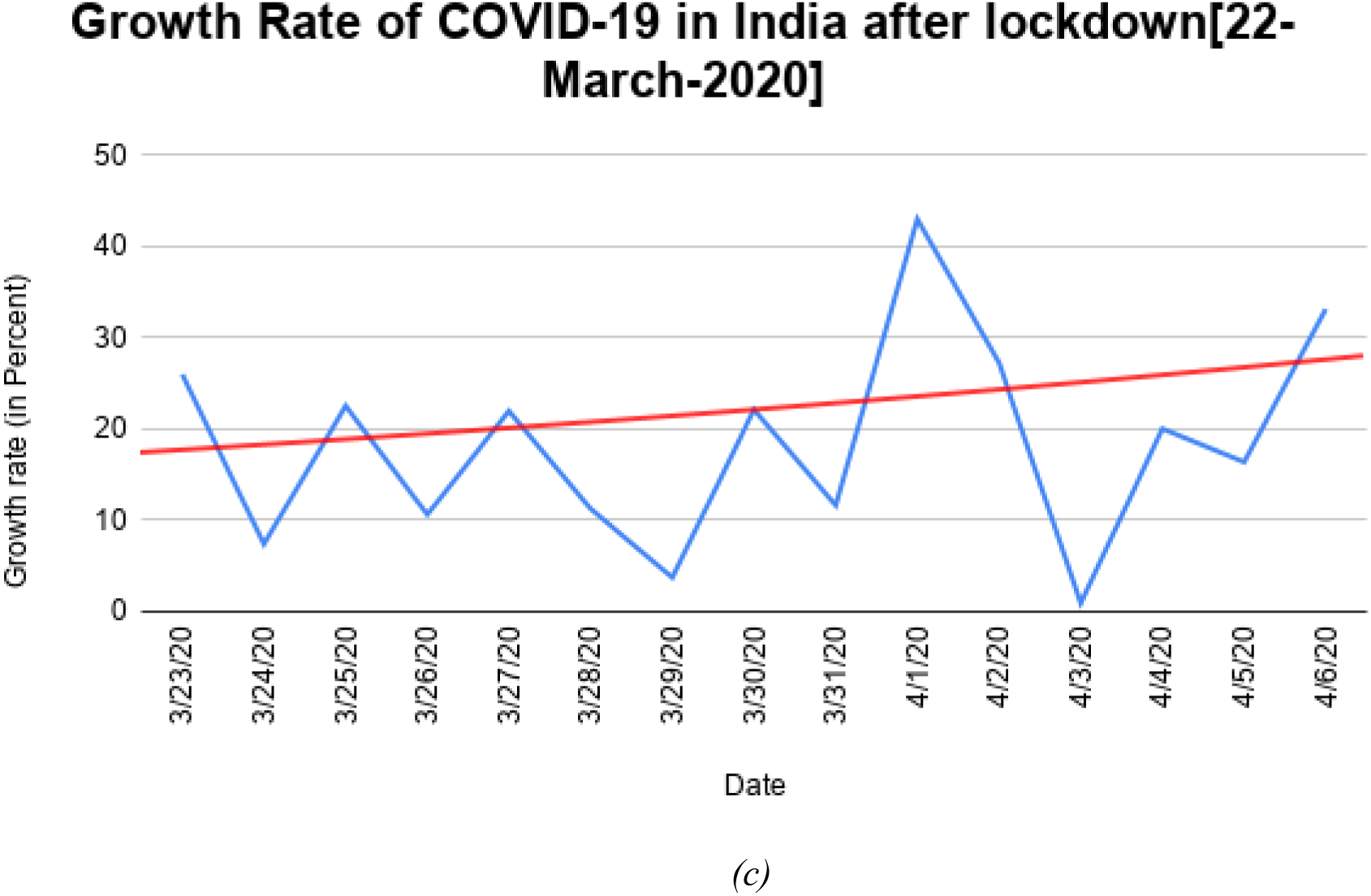
Graphs showing the growth rate of infected cases on daily basis (blue line) and trend line of expected growth rate (red line) divided into three phases (a) Early Phase: Time Period between 22^nd^ January to 4^th^ March 2020 (b) Prior to lockdown period: Time Period between 5^th^ March 2020 to 22^nd^ March 2020 (c) Post Lockdown period: Time Period after 22^nd^ March 2020 to 6^th^ April 2020

### b. Short term predictions for Infected cases

Exponential Modelling has been used to predict short term predictions at national level. Firstly, the growth for doubling days was calculated, i.e. the number of days to double the number of infected cases has been calculated. As seen from Figure 4, the first image shows that prior to the lockdown period average number of days to double the cases was majorly above four, while the average period drop down near to three after lockdown. As per the predictions (trend line in red) the average number of days is constantly decreasing with the rising number of cases in India.

**Figure 4.**
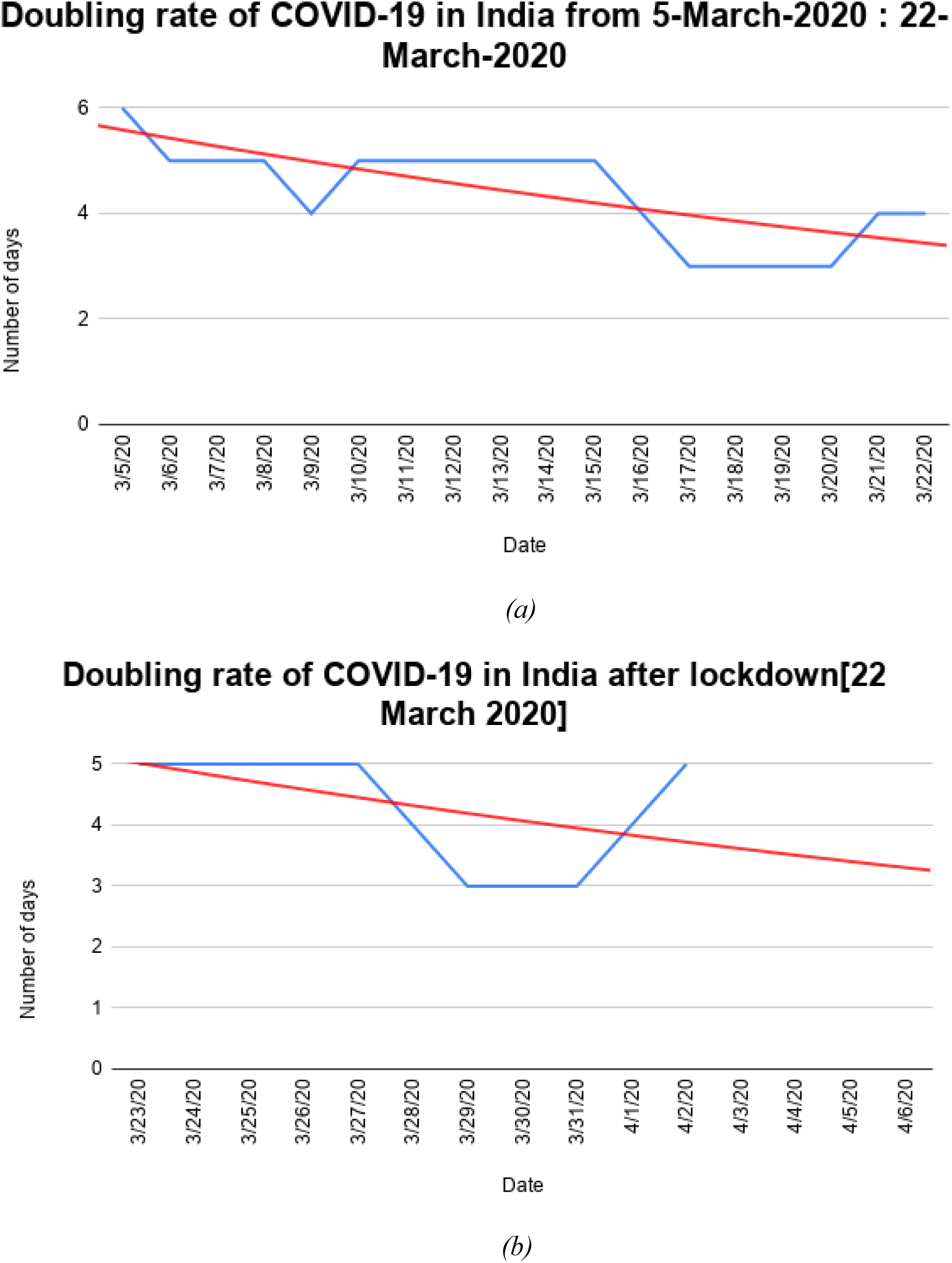
Doubling rate of number of days for the number of infected cases to double in India as per the Exponential modelling, with blue line denoting the actual rate while red line denoting the predictive line (a) Doubling rate before the lockdown period i.e. 22^nd^ March 2020 (b) Doubling rate after the lockdown period i.e. 22^nd^ March 2020

Based on the exponential model, the predictions for the next 3 weeks were made for the infected cases in India. Considering that doubling rate is going as per the historical evidences, the number of predicted cases in India is shown in Table 1. Based on the exponential modelling based growth of the number of COVID-19 cases in India, polynomial regression line was plotted with different degree values. A total of 5 degrees were checked between 2 to 6 and Root Mean Error (RME) was checked for all cases. The lowest RME reported was 237.58 for the model with degree 4. Thereafter, the prediction model of polynomial regression of degree 4 was built and used for the prediction values. For training set, data for 31 days was considered starting from 3-March-2020 to 3-April-2020. This was due to the fact that prior to 3^rd^ March, only 5 cases were reported in India within 40 days, which was impacting the prediction model. For evaluation, data from a short time period of 4-April-2020 to 7-April-2020 was used. Other combinations of training and test sets were also considered, but the predictions were similar. With these prediction values, it is estimated that the values for infected cases may rise near to 75,000 in India by end of current month, which may not be a really good situation in India.

**Table 1.** Predicted number of Infected Cases in India based on Exponential Modelling for next few days.

| Date | Predicted Number of Infected Cases |
| --- | --- |
| 4/07/2020 | 5245 |
| 4/08/2020 | 6111 |
| 4/09/2020 | 7088 |
| 4/10/2020 | 8187 |
| 4/11/2020 | 9416 |
| 4/12/2020 | 10786 |
| 4/13/2020 | 12308 |
| 4/14/2020 | 13993 |
| 4/15/2020 | 15851 |
| 4/16/2020 | 17895 |
| 4/17/2020 | 20137 |
| 4/18/2020 | 22589 |
| 4/19/2020 | 25264 |
| 4/20/2020 | 28175 |
| 4/21/2020 | 31336 |
| 4/22/2020 | 34761 |
| 4/23/2020 | 38464 |
| 4/24/2020 | 42460 |
| 4/25/2020 | 46763 |
| 4/26/2020 | 51391 |
| 4/27/2020 | 56358 |
| 4/28/2020 | 61680 |
| 4/29/2020 | 67375 |
| 4/30/2020 | 73458 |
| 5/01/2020 | 79949 |
| 5/02/2020 | 86864 |

### c. Social Distancing

The concept of social distancing was suggested by a lot of health experts around the world as it was important that the chain of physical interaction between humans must be broken. The major step in this direction was to announce lockdown at national level. Indian government took this suggestion seriously and started with the one-day implementation of lockdown on 22^nd^ March 2020 and then announcing a 21-day lockdown till 14^th^ April 2020. It was welcomed by a lot of citizens in India. The same was suggested by Google through its mobility report as shown in Figure 5.

**Figure 5.**
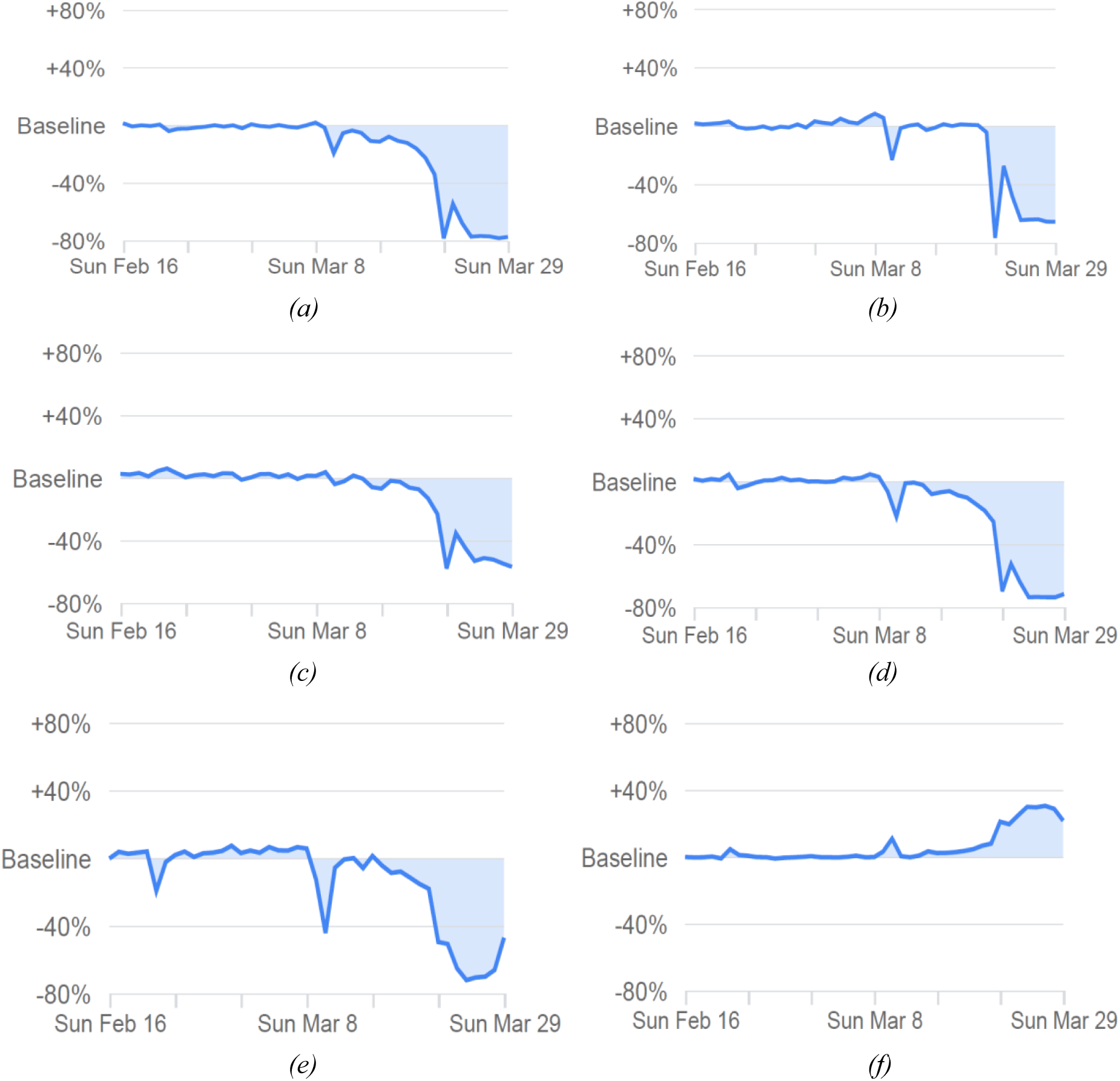
Mobility changes in India till 29^th^ March 2020 (a) Graph showing 77% decline as compared to the baseline in the Retail and recreation places like restaurants, cafes, shopping centers, theme parks, movie theatres, etc. (b) Graph showing 65% decline as compared to the baseline in the Grocery & pharmacy like markets, food shops, farmer places, drug stores, pharmacies, etc. (c) Graph showing 57% decline as compared to the baseline in the Parks like public beaches, dog parks, marinas, national gardens, public gardens, etc. (d) Graph showing 71% decline as compared to the baseline in the transit stations like bus stations, public transports, metro stations, train stations, etc. (e) Graph showing 47% decline as compared to the baseline at the Workplaces like private and government offices (f) Graph showing 22% increase as compared to the baseline at the residential places (Source: Google Mobility Trends Report)

As per the Google’s method of calculation, every day changes for the number of visits and length of stay at different places were compared the baseline values. The baseline value was considered as the median value for the corresponding day of the week. There were six different categories which were considered important for social distancing and there was a significant drop down seen in five of these categories. In Figure 5, Graph shows a 77% decline as compared to the baseline in the visits at Retail and recreation places like restaurants, cafes, shopping centers, theme parks, movie theatres, etc.; 65% decline as compared to the baseline in the visits to the Grocery & pharmacy places like markets, food shops, farmer places, drug stores, pharmacies, etc.; 57% decline as compared to the baseline in the visits to the Parks like public beaches, dog parks, marinas, national gardens, public gardens, etc.; 71% decline as compared to the baseline in the transit stations like bus stations, public transports, metro stations, train stations, etc.; and 47% decline as compared to the baseline at the Workplaces like private and government offices. It was seen that a 22% increase as compared to the baseline was observed at the residential places which implies that more Indians are now staying at their homes. All these numbers cannot be 100% as a lot of frontline health workers, administrators, pharmacists, grocery store owners and people involved in other essential services were on duty during the lockdown as well. However, the basic purpose of lockdown to maintain social distancing has been achieved to good potential. Citizens are spending more time at their homes as compared to crowded places. So it can be said that Indian citizens have been able to follow the lockdown effectively.

### d. Impact of Mass Events on infected cases

During the lockdown period, there were two mass events reported in India. One was related to the exodus of the laborers to their respective states in India [23] and the other was a religious event which happened in New Delhi at a mass level [24]. There are around 25% citizens living below poverty line and have to depend on the daily wages to feed their families. Once the lockdown was announced, the fate of these 1.3 billion people was under scanner and that is why even the Government of India came up with a package of more than 22 billion USD to help these workers and laborers. All the respective state governments also came up with the different infrastructural setup for providing food and money to the needy citizens. Some agencies reported that in providing food security, India and the neighboring nations depending on India may fall short of food [25] while the others reported loss of millions of job [26] during the lockdown due to such mass exodus. However, the numbers of infected cases were not impacted much by this mass movement as majority of workers were not carrying any infection with them during their movements from their workplaces regions to their native place region.

But there was a serious impact seen on infected cases due to conduction of a religious event in Delhi. As seen from Figure 6, the average number of days to double the infected cases from corona virus without any cluster event was estimated to be 7.1 as per the health ministry, while it was 4.1 after Delhi’s religious event took place [27]. This event resulted in formation of clusters in the whole country as many people who attended this event went to different parts of the country without following any rules of getting quarantined. Some of them who came from out of India to attend this event did not follow rules and advisories issued regarding COVID-19 protection. This resulted in a sharp spike in the number of infected cases after 31^st^ March 2020.

**Figure 6.**
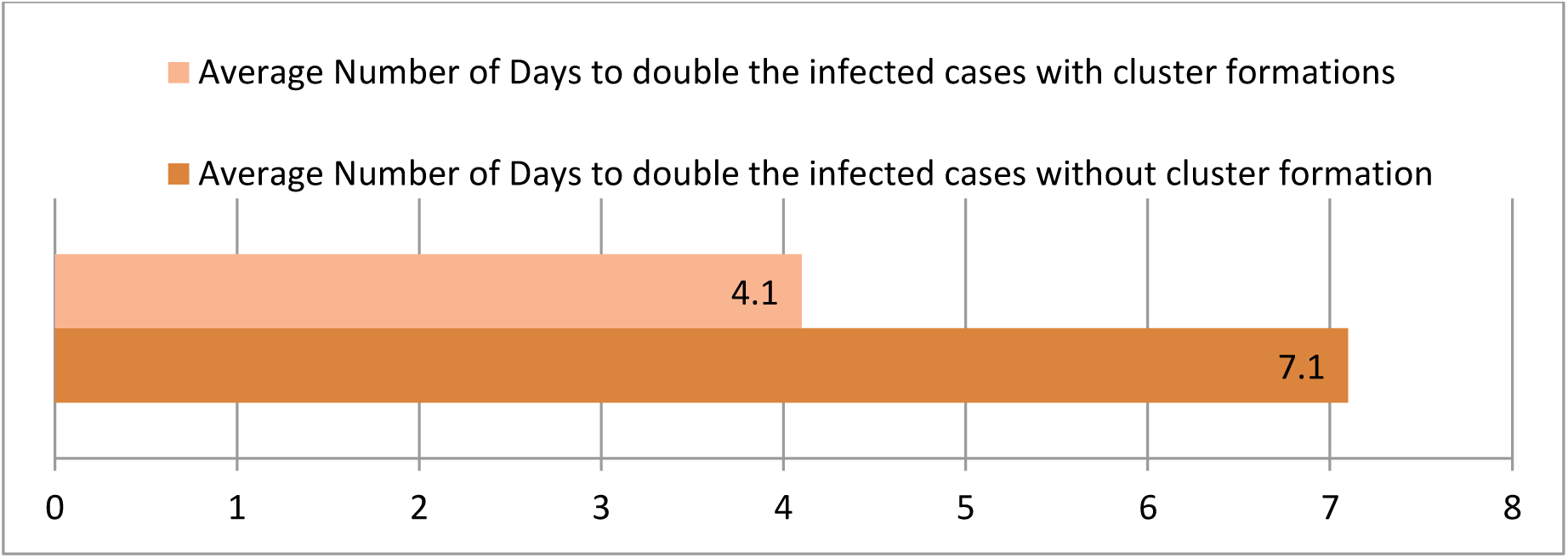
Graph showing the average number of days for the infected cases to double with or without cluster events.

As per the Figure 7, at least 36% of the total infected cases reported on 6^th^ April 2020 were linked to the religious event in Delhi [28]. Out of over 4000 cases reported, a minimum of 1445 cases were suspected to be due to the religious event in Delhi. This event took the graph sky high in a week’s time with growth slowly moving towards exponential path and India entering into the third phase of community transmission for COVID-19 virus. Many regions of India which were totally isolated with the infection of Corona Virus, also reported their first case in the first 3-4 days of the conduction of the event. Therefore, it can be said that religious event that happened in Delhi has really pushed the bars high for the number of infected cases from coronavirus in India. It has far more serious consequences than it seems right now and authorities needs to be really alert as people linked to this event have gone back to different parts of the nation.

**Figure 7.**
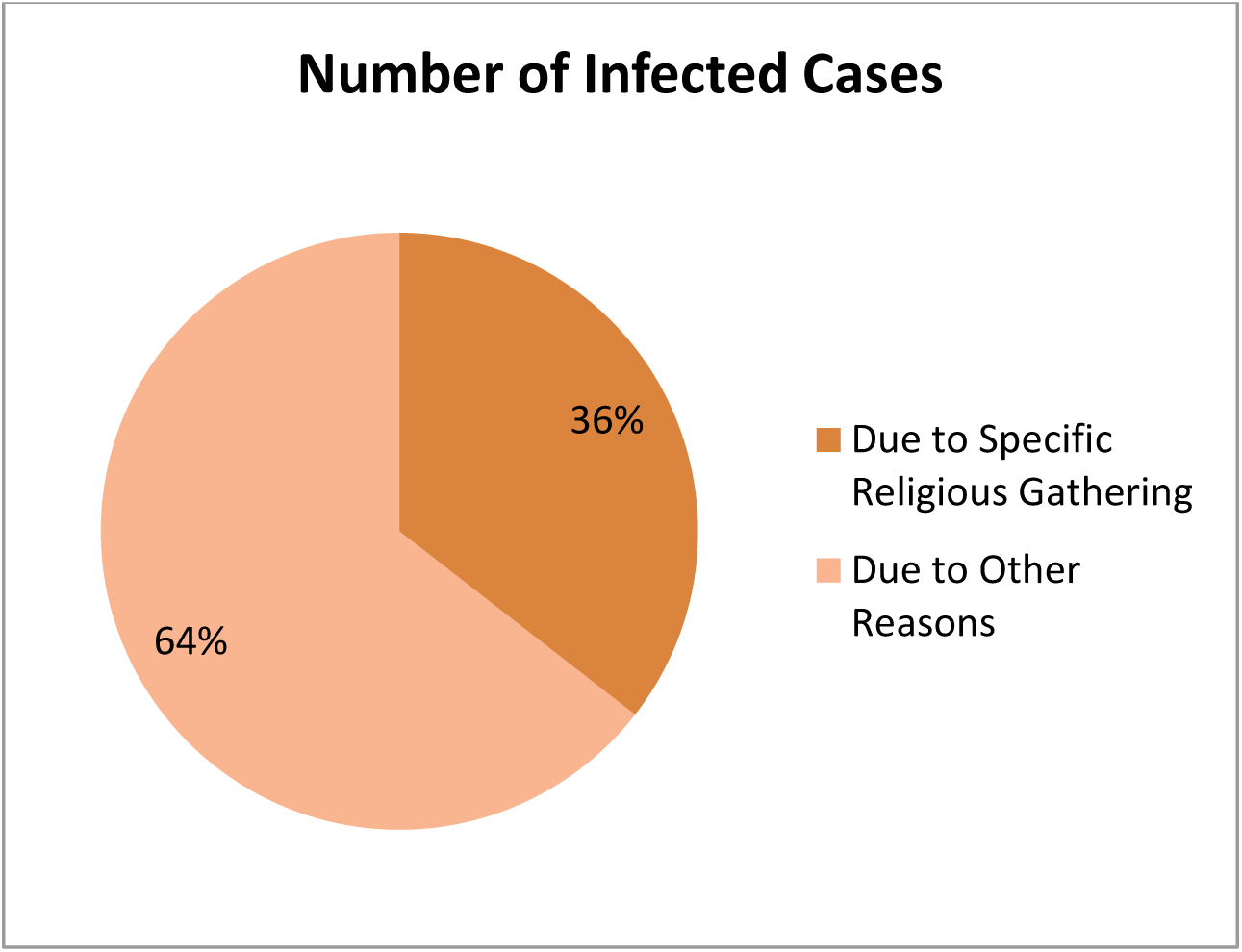
Percentage distribution of Infected Cases due to religious event in Delhi out of the total infected cases as per the data released by Ministry of Health, Government of India

### e. Network Analysis & Classification of Infected patients

Based on the data available from crowdsourced database at COVID-19-India dot org [17], network of the patients and their demographic details was created. The patient ID, the countries of travel, and any mass events were considered as the nodes and connection between each patient and their travelling history or event attending history was considered as edge in the network. Presenting the visualization of the network consisting of 551 valid nodes was out of scope for this paper; however degree centrality of important nodes was calculated and is presented in Table 2. Degree centrality of the nodes is calculated as the number of connections with that particular node divided by the total number of edges present in the network. The top 7 nodes based on their nodes centralities were Religious event in Delhi, Italy, Gulf Countries, United Kingdom, Mumbai, Saudi Arabia and the first patient. It was found that these were the major hotspots responsible for the rapid spread of coronavirus in India.

**Table 2.** Degree Centrality Measure for top seven nodes in the network.

| Node Description | Degree Centrality Measure |
| --- | --- |
| Religious Event in Delhi | 0.102775941 |
| Italy | 0.030072703 |
| Gulf Countries | 0.011566642 |
| United Kingdom | 0.007270324 |
| Mumbai | 0.005948447 |
| Saudi Arabia | 0.004296100 |
| First Patient | 0.000370921 |

Based on the patient database, classification model was developed to check that on the basis of the demographic features, can a patient have the probability to be deceased. Other patterns were mined for the rules for COVID-19 patients; however rules were not significant enough for the positive cases as data for negative cases was not available in abundance. Based on the age group of COVID-19 infected patients, it was found that majority cases were in the age group of 31-40 years in India, as shown in Figure 8. The deceased patients were majorly above 60 years of age group and majority people who are infected either travelled back from Italy or attended the religious event in Delhi.

**Figure 8.**
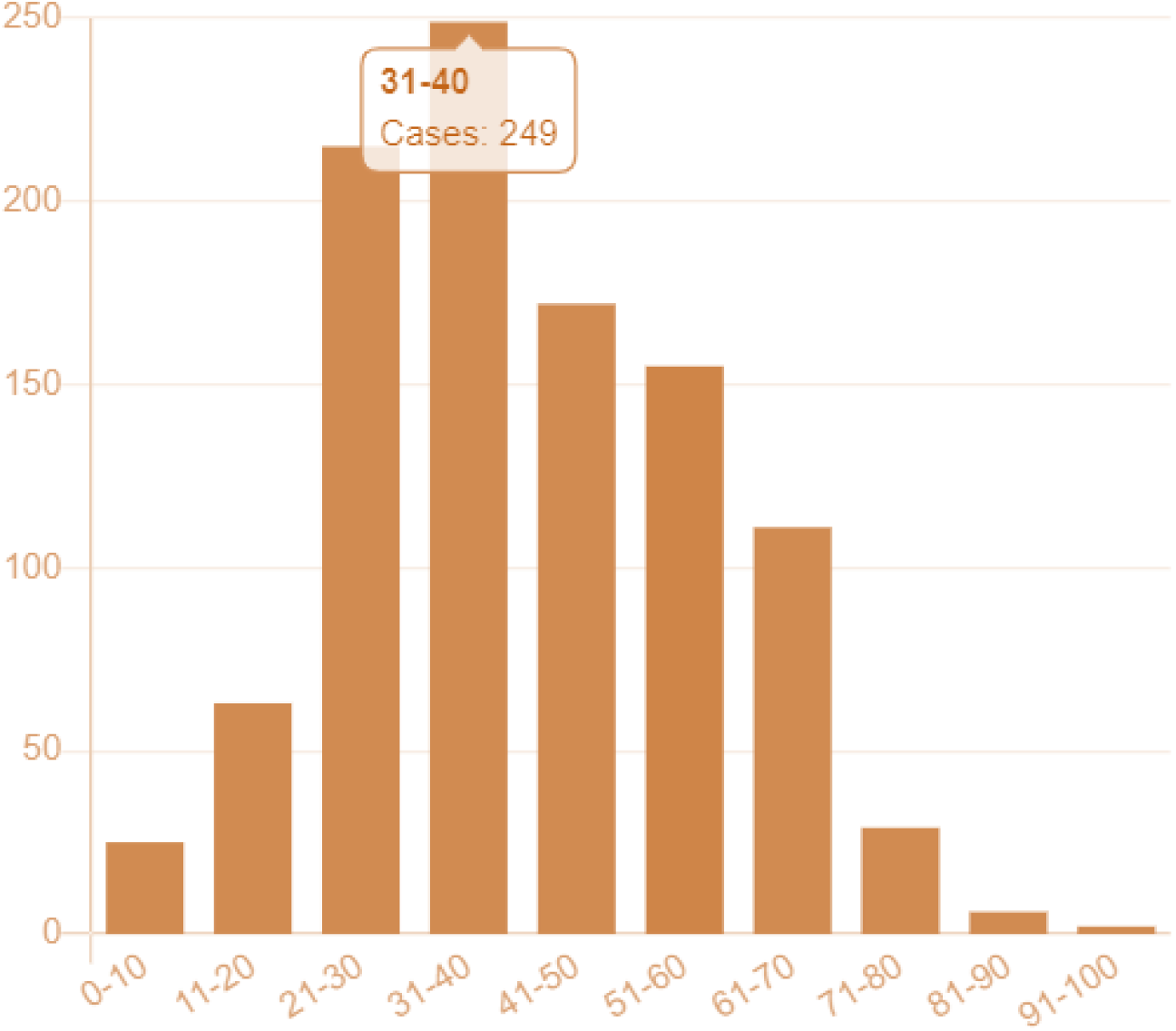
Age distribution of patients infected from COVID-19 [17].

There was a classification model developed around patients who lost their lives after getting infected from the coronavirus. Decision Tree classification model [29] was used to classify that out of the given COVID-19 infected patients, whether a patient will be deceased or not. Three features were found to be significant which were the Age of the Patient, Gender of the patient, and the state/region of the patient. Although a total of 5000 patients have been infected from COVID-19 virus so far, but demographic details of all the patients is not known.

There were 912 living patients and 129 dead patients modeled from the dataset. Out of 129, only 24 patients had complete demographic details available for the modelling, however for rest of the patients, relevant data imputation techniques [30] were used and missing values were filled up scientifically. Accuracy and Precision values were computed based on the confusion matrix generated from Decision Tree Classifier model. The model generated a 60% accuracy as the data points were too less for the model to be trained and tested. Moreover, the three-variable model was not sufficient enough to make any significant relevance out of the analysis. However, the deaths have found to be more prevalent in the age group of 60 or more years and Males were more prominent to getting deceased. Also, infants were least impacted by coronavirus in terms of life and death. The state of Maharashtra and Madhya Pradesh were found to be the significant regions for the dead cases. Not major inferences could be drawn from the classification model as numbers of cases were low, but as per the records of deceased patients, a hint can be taken by the medical and administrative authorities as in which type of patients (based on their demographical features) need extra critical care.

### f. Future Plans for Lockdown and other strategies

Considering the numbers in previous sections, there are five major reasons due to which the national lockdown will be difficult to be abolished. Firstly, the growth rate of infected cases is continuously increasing. Secondly, the doubling time for number of infected cases in India is declining very fast making it a dynamic spread. Thirdly, majority of the Indians are cooperating and following the social distancing fairly well with mobility rates going down for public places in India. This is due to the combined efforts of the administrative authorities, engagement activities by the office of Prime Minister and citizen’s will to stay away from social events. Fourthly, the sudden mass events are ruining the efforts of the Indian authorities to contain the spread of Coronavirus. And lastly, the penetration of the people infected from religious event has been really deep in India currently. There is a lot of tension created in India due to spread of infected people and the numbers are rising with each day. Thus, it is very difficult for India to completely ban the National Lockdown after 14^th^ April.

In extreme conditions, the state wise rate of infection spread needs to be considered and then decision over lifting the lockdown may be taken. Table 3 shows the current number of infected cases in different states of India and the associated growth rates for the last 7 days for each state, respectively [17]. It can be seen that majority of the states have witnessed more than 200% of growth rate in last one week, which is worrisome situation for all the state authorities. In fact, the national growth has also been towards 180% mark which is very high number is in the last seven days. The administrators for the regions with less than 50 infected cases can consider the uplifting of ban on lockdown in their respective regions, however uplifting national lockdown looks difficult. The social distancing must be followed for next few weeks so that curve flattening process can continue for some more time.

**Table 3.** Number of Infected Cases and last 1-week growth rates for different states of India.

| States | Number of Infected Cases<br>as on 8<sup>th</sup> April 2020 | 1-Week Growth Rate<br>(1<sup>st</sup> April – 7<sup>th</sup> April'20) |
| --- | --- | --- |
| MAHARASHTRA | 1135 | 238.81 |
| TAMIL NADU | 738 | 215.38 |
| DELHI | 576 | 278.95 |
| TELANGANA | 453 | 256.69 |
| RAJASTHAN | 363 | 202.50 |
| UTTAR PRADESH | 361 | 208.55 |
| ANDHRA PRADESH | 348 | 213.51 |
| KERALA | 345 | 30.19 |
| MADHYA PRADESH | 290 | 195.92 |
| GUJARAT | 186 | 126.83 |
| KARNATAKA | 181 | 64.55 |
| HARYANA | 167 | 288.37 |
| JAMMU AND KASHMIR | 158 | 154.84 |
| PUNJAB | 106 | 140.91 |
| WEST BENGAL | 99 | 266.67 |
| ODISHA | 42 | 950.00 |
| BIHAR | 38 | 65.22 |
| UTTARAKHAND | 33 | 371.43 |
| ASSAM | 28 | 75.00 |
| HIMACHAL PRADESH | 27 | 575.00 |
| CHANDIGARH | 18 | 38.46 |
| LADAKH | 14 | 7.69 |
| ANDAMAN AND NICOBAR ISLANDS | 11 | 10.00 |
| CHHATTISGARH | 10 | -44.44 |
| GOA | 7 | 16.67 |
| PUDUCHERRY | 5 | 66.67 |
| JHARKHAND | 4 | 300.00 |
| MANIPUR | 2 | 100.00 |
| ARUNACHAL PRADESH | 1 | NA |
| DADRA AND NAGAR HAVELI | 1 | NA |
| MIZORAM | 1 | NA |
| TRIPURA | 1 | NA |
| TOTAL | 5749 | 180.85 |

With respect to transportation services, only intrastate travel may be allowed in various states of India where population density is low and reported cases are less than 50 right now. No inter-state domestic travel should be allowed for the time being as it may result in transferring the COVID-19 carriers from one region to another region. International travel flights must not be activated as the number of cases in majority of the regions of the world is very high. European and US regions are already facing most of the turmoil, while other parts of the world are seeing rapid rise in number of infected cases. Starting inbound international flights to India may disturb the COVID-19 action plan in India. Only essential services should be open for the citizens of India and lockdown should be carried on for next 2-4 weeks.

Due to expected shortage of doctors, health workers and essential service providers, the Government of India should rope in the military, BSF, corporate, NGOs, students and volunteers. Provisions should be made in parallel to train them online and prepare them for the worst. Meanwhile, the Government of India should also plan to arrange Personal Protective Equipment & other resources for their optimal participation in the national mission. Lockdown with more stringent controls should be enforced for the next few weeks by identifying hotspots and isolating them. Due to the population density of the country, customized region wise solutions should be exercised with relaxation to practice livelihood in safer identified regions. For this, central level administrative unit, state level administrative unit and local level administrative units need to communicate with each other efficiently. The disease can be better controlled in India due to massive stock of suitable medication available in the country, which is why India is also able to support other nations in terms of drug supply.

## 5. Conclusion

This study presented a comprehensive analysis of the COVID-19 outbreak situation in India. The cases are rising very fast and they need aggressive control strategies from the Administrative units of India. There are six different aspects covered up in this study and six research questions have been answered comprehensively. They are related to presenting the growth trends of infected cases in India, predictions for the number of infected cases for next few days, impact of social distancing on the citizens of India, impact of mass events on the number of infected cases in India, network analysis and mining of patterns on the patients suffering from coronavirus, and analyzing the strategies for uplifting lockdown in India. The current study implemented various techniques to present the data analysis and the results are in sync with few limited studies available in the literature. This study will be useful for the Government of India and various states of India, Administrative Units of India, Frontline health workforce of India, researchers and scientists. This study will also be favorable for the administrative units of other countries to consider various aspects related to the control of COVID-19 outspread in their respective regions.

## Data Availability

Data is available publicly from John Hopkins University repository and COVID-19-INDIA dot org.

## Acknowledgement

Authors would like to acknowledge the continuous efforts of Ministry of Health and Family Welfare, Government of India (https://www.mohfw.gov.in/) and different state units for providing updated numbers for COVID-19 cases in India through their portal and press releases. The authors would also like to acknowledge the exhaustive data collation work done by the team of COVID-19-India dot org (https://www.covid19india.org/).

## About Authors

**Dr. Rajan Gupta** is a Research and Analytics professional and is currently associated with University of Delhi in India as Assistant Professor. He has done his PhD from University of Delhi in the area of Application of Data Science in Improving E-Governance in Developing Nations. He has written two books on E-Governance in India and has published various papers at national and international forums. He is an active consultant to a lot of organizations in the area of Analytics, Bio-statistics and Data Science.

**Dr. Saibal K Pal** is from DRDO, a R&D organization under the Ministry of Defense, Government of India. He has done his PhD from University of Delhi and has been involved in lot of projects related to implementation of E-Governance in India. His area of research includes security, information systems and E-Governance. He has published two books on E-Governance in India and has published various papers at national and international forums.

**Mr. Gaurav Pandey** is a research student at TheNorthCap University Gurugram and is also currently an intern at ISRO office. His areas of interest are Data Science, Machine Learning and Artificial Intelligence using Python.

